# Multitissue Multiomics Integration Reveals Tissue-Specific Pathways, Gene Networks, and Drug Candidates for Type 1 Diabetes

**DOI:** 10.1101/2024.11.25.24317912

**Authors:** Montgomery Blencowe, Zara Saleem, Ruoshui Liu, I-Hsin Tseng, Julian Wier, Xia Yang

## Abstract

**Aims/hypothesis:** Although genome-wide association studies (GWAS) have identified loci associated with Type 1 diabetes (T1D), the specific pathways and regulatory networks linking these loci to disease pathology remain largely unknown. We hypothesized that T1D genetic risk factors disrupt tissue-specific biological pathways and gene networks that ultimately lead to beta cell loss.

**Methods:** We conducted a multitissue multiomics analysis that integrates human GWAS data for T1D with tissue-specific regulatory data on gene expression and gene network models across relevant tissues to highlight key pathways and key driver genes contributing to T1D pathogenesis. Key driver genes were validated using islet-specific gene expression and protein data from non-obese diabetic (NOD) mice compared to non-T1D mouse models. Drug repositioning predictions were generated using the L1000 and PharmOmics platforms.

**Results:** Our integrative genomics approach identified known immune pathways across multiple tissues, such as adaptive immune responses, cytokine-mediated inflammation, primary immunodeficiency, and interactions between lymphoid and non-lymphoid cells. Tissue-specific signals included genes related to type 2 diabetes in lymphocytes, viral response pathways in macrophages and monocytes, and Notch signaling in adipose and immune cells. In pancreatic islet analysis, we observed significant enrichment for T1D and type 2 diabetes gene sets alongside immune-related pathways, including antigen processing, systemic lupus erythematosus, and interferon signaling. Removing HLA genes from the analysis revealed additional immune pathways, such as RIG-I/MDA5 induction of interferons, along with melanogenesis, steroid hormone synthesis, and iron transport. Network modeling highlighted the autoimmune basis of disease with key drivers such as FYN, TAP1, WAS, and HLA-B/C/G, as well as further immunomodulatory genes such as LCK, LCP2, EMR1, and GC. These key drivers were further supported by gene and protein expression data from NOD mice. We additionally highlight various drug classes that target the T1D genetic networks and may be useful to delay T1D development.

**Conclusions/interpretation:** Our multitissue multiomics approach provides a detailed landscape of the tissue-specific genetic networks and regulators underlying T1D. This analysis confirms the roles of known immune pathways while uncovering additional regulatory elements and disease-associated networks, thus expanding our understanding of T1D pathogenesis. The identification of potential drug candidates through network analysis offers potential therapeutic strategies for targeting disease pathways and holds promise for delaying or preventing T1D progression.

## INTRODUCTION

Type 1 diabetes (T1D) is characterized as the autoimmune loss of pancreatic beta cells and resultant impairment of glucose homeostasis. Currently, T1D affects ∼8.5 million people globally, representing 5-10% of the total diabetic population with an annual incidence increase of 2-3%. (1–3). Risk of developing T1D is increased by approximately 5.6% and 50% with an affected parent or diseased monozygotic twin, respectively, when compared to the general population. Parental heritability estimates predict that diabetic fathers confer an increased risk of T1D development of about 12% while mothers at around 6% (4). Thus, there is a genetic component predisposing T1D pathogenesis. However, T1D is not usually present in individuals with a family history, with only around 10% of patients having a first/second-degree relative with the disease, thus also suggesting a significant environmental contribution (5). Interestingly, there seems to be an increased disposition for developing T1D if one lives in regions of Northern Europe, independent of genetic background, highlighted by an increase in T1D incidence for migrants living within these regions (6,7). This environmental contribution has several potential manifestations such as alterations in gut microbiota (8) or pre- and post-natal dietary factors, including early exposure to gluten (9) and Vitamin D deficiency (10). Furthermore, a longstanding hypothesis predicts that the exposure to viral infection may also be a causal factor (11), particularly enteroviruses (12) which seem to target pancreatic islet cells (13). Therefore, both genetic and environmental components contribute to T1D incidence and progression, yet a large gap exists in understanding the complex genetic and environmental architectures as well as the interaction between the two. It is plausible that genetic risks represent first hits, and environmental factors act as second hits to interact with genetic predisposition to trigger T1D development (14–16).

On the genetic front, genome wide association studies (GWAS) have uncovered over 60 T1D genetic risk loci. The main genes predisposing T1D patients are located within the HLA region on chromosome 6, encoding the major histocompatibility complex (MHC), which is critical for adaptive immunity. While HLA-encoding genes have the strongest association and account for up to 50% of the total genetic T1D risk (17,18), loci outside of the HLA region including protein tyrosine phosphatase, non-receptor type 22 (*PTPN22*), interleukin 2 receptor alpha (*IL2RA*), insulin gene (*INS*), and cytotoxic T-lymphocyte-associated protein 4 (*CTLA4*), have also been associated with disease development (19). However, these top loci at genome-wide significance cannot fully explain the total genetic heritability of T1D. Moreover, the dominance of the HLA effect may overshadow additional unknown and important processes contributing to T1D pathogenesis with or without interactions with environmental factors. Identifying the missing genetic risks, or the “dark matter”, is important to gain a full understanding of the genetic underpinnings of T1D pathogenesis. In addition, growing lines of evidence support an “omnigenic” disease model (20), which states that a large proportion of genes of the genome may contribute to disease development through gene-gene interactions in networks within and between tissues, and key network regulators likely play more central roles than other peripheral disease associated genes in the networks. In support of this, top GWAS loci for complex diseases have been found to be more concentrated in the periphery of gene networks and are less likely to be network regulators. Therefore, simply focusing on the top GWAS hits will likely miss crucial regulatory genes. We hypothesized that T1D genetic risks with a wide spectrum of effect sizes (strong, moderate, or subtle) interact and perturb tissue-specific gene networks through a select set of regulatory genes, resulting in variations in T1D susceptibility.

Integration of GWAS data with functional genomics information such as tissue-specific expression quantitative trait loci (eQTLs) and gene networks has proven to be a powerful tool to pinpoint causal genes and their associated pathogenic mechanisms and regulators within the context of specific tissues (21–32). In this study, we use a computational pipeline, Mergeomics (**Figure 1**) (25,30) to integrate T1D GWAS studies with tissue-specific functional information, such as genetics of gene expression and gene regulatory networks from a broad range of T1D relevant tissues or cell types including blood, lymphocyte, macrophage, monocyte, pancreas, islet, subcutaneous adipose, and visceral omentum adipose (33–42). Our analysis not only confirmed the importance of immune pathways across various tissues, but revealed numerous tissue-specific pathways, networks and regulators. By understanding how T1D genetic risks affect gene networks within and across tissues and identifying causal key regulators, our study offers comprehensive insights into T1D pathogenesis and helps prioritize regulators and potential therapeutic strategies.

**Figure 1:**
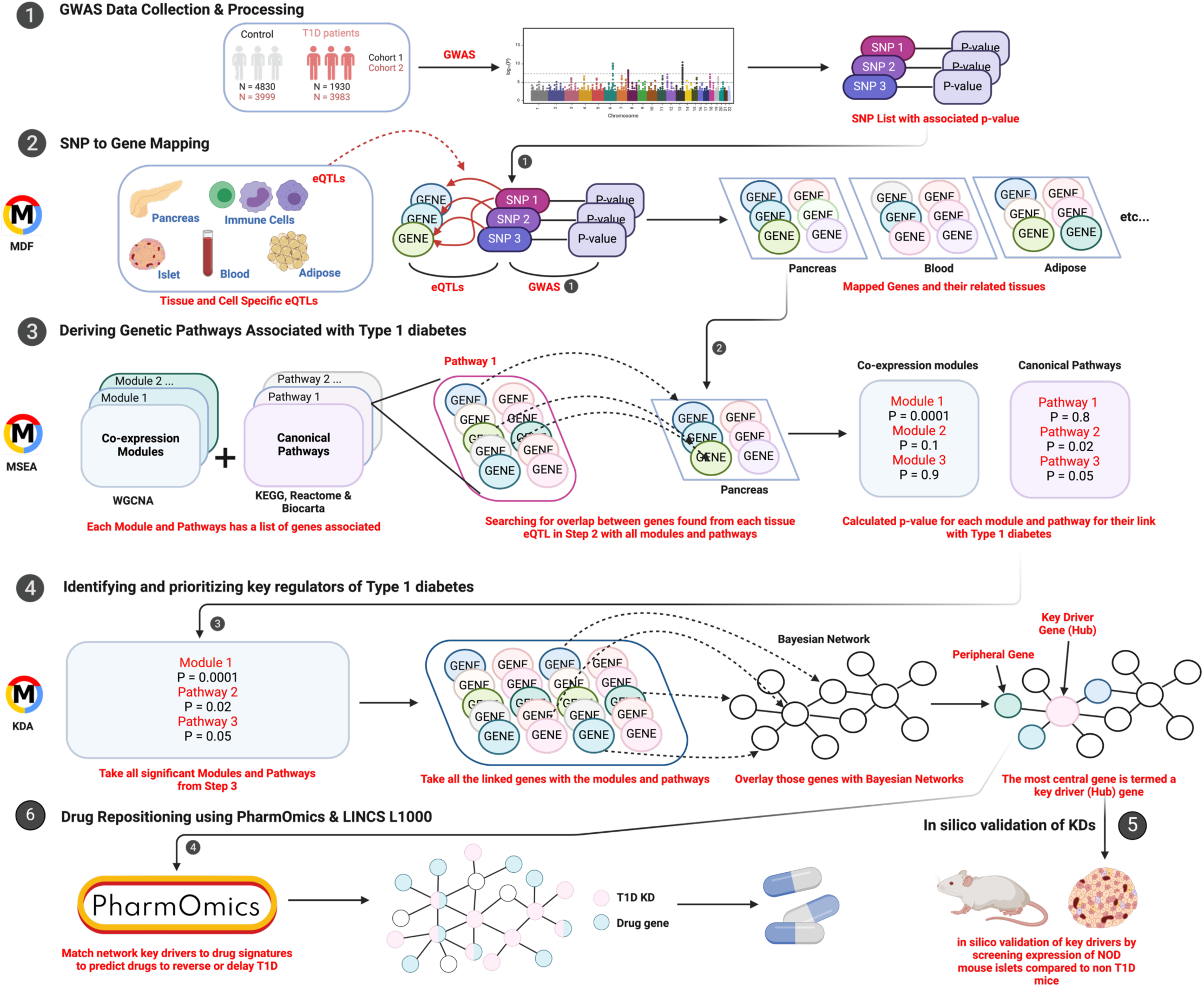
Overview of Study. We integrated the following datasets using Mergeomics: 1) Collect and process T1D GWAS SNPs. 2) Map SNP to genes using tissue specific eQTLs. 3) Genes found from step 2 are then linked to canonical pathways and co-expression modules. 4) Implement wKDA using both protein-protein interaction (PPI) and Bayesian networks independently for key driver gene identification. 5) Conduct *in silico* validation of key drivers. 6) Perform drug repositioning using PharmOmics and LINCS L1000.

## RESULTS

### Overview of study design

As depicted in Figure 1, we first assessed which biological pathways were enriched for T1D GWAS signals based on GWAS data from the JDRF/Wellcome Diabetes and Inflammation Laboratory at the University of Oxford (43). The T1D and control individuals were partitioned into two independent cohorts based on the genotyping platforms (i.e., Illumina or Affymetrix) between cases and controls. Cohort 1 was comprised of 1930 T1D patients and 4830 Controls, and the Affymetrix platform was used. Cohort 2 had 3983 T1D patients and 3999 Controls, and the Illumina platform was used. Standard GWAS analysis was carried out for each cohort separately and we used the full GWAS summary statistics from these two independent cohorts.

To identify tissue-specific pathways associated with T1D GWAS single nucleotide polymorphisms (SNPs), we utilized tissue-specific eQTLs to guide SNP-to-gene mapping. This analysis included a total of 18 eQTL sets, with 13 from the GTEx database (44,45) covering subcutaneous adipose, visceral omentum adipose, blood, brain, colon, heart, liver, lymphocyte, muscle, pancreas, pituitary, spleen, and stomach tissues, along with macrophage and monocyte eQTL sets from the Cardiogenics Consortium (29), immune cell eQTLs (including lymphocytes) from the DICE study (46), and pancreatic islet-specific eQTLs (5). The genes mapped to the GWAS SNPs through each tissue-specific eQTL set were analyzed against biological pathways based on literature-based functional categories and tissue-specific gene coexpression network modules which define data-driven functional genes sets using Marker Dependency Filtering (MDF) and Marker Set Enrichment Analysis (MSEA) from Mergeomics (25,30). The literature-driven pathways were taken from Reactome (Version 45) (23), Biocarta (47), and the Kyoto Encyclopedia of Genes and Genomes (KEGG) databases (48). The gene coexpression modules were derived using the Weighted Gene Correlation Network Analysis (WGCNA) approach (49) to construct networks from tissue-specific transcriptomics datasets from GTEx (44). This approach identifies functionally related gene sets based on gene expression patterns and has previously helped derive novel biological insights into various diseases (26–28,31,50–52). Integration of T1D GWAS, tissue-specific eQTLs, and pathways and coexpression modules together using MSEA reveal pathways and modules enriched for stronger genetic associations with T1D compared to random gene sets. Following MSEA, we next carried out a Meta-MSEA to meta-analyze the two independent GWAS datasets to look for shared pathways/modules, which we further simplified into independent “supersets” to reduce redundancy between similar pathways/modules. Integrating these supersets with gene regulatory networks (GRNs) and protein-protein interaction networks (PPI), we carried out a key driver analysis (KDA) to identify key drivers (KDs), which are central network genes whose network neighbors are highly enriched for genes in the T1D pathways/modules. These KDs were then visualized in tissue-specific networks. We further carried out *in silico* validation using curated islet gene expression and protein data from eight strains of mice to highlight if our KDs were significantly and uniquely changing in a T1D mouse model. Lastly, we used our KDs as input into L1000 and PharmOmics to highlight potential drug candidates for T1D treatment.

### Identification of consistent and divergent canonical pathways and gene co-expression modules associated with T1D across cohorts and tissues

We assessed which knowledge-based biological (canonical) pathways and data-driven gene co-expression modules were enriched for T1D GWAS signals. The use of tissue-specific eQTLs served to guide SNP-to-gene mapping, allowing us to capture tissue specific results. From the 18 eQTLs sets used, we found the following seven tissues to be the most informative in terms of how many significant pathways were identified: blood, lymphocyte, macrophage, monocyte, pancreas, subcutaneous adipose, and visceral omentum adipose. We therefore focused on reporting the results from these seven informative tissues only, with other tissues serving to supplement the main results.

Out of the 1827 curated canonical pathways, we identified 187 pathways enriched for T1D GWAS association from Cohort 1 at an FDR < 5% in at least one of the seven chosen tissues (blood, macrophage, monocyte, lymphocyte, pancreas, subcutaneous adipose and visceral adipose; **Supplement Table 1A**). From Cohort 2 we found a total of 143 pathways enriched for T1D GWAS association at an FDR < 5% (**Supplement Table 1B**). Between Cohort 1 and Cohort 2, we found 121 pathways significant in both, 66 unique to Cohort 1 and 22 unique to Cohort 2 (**Figure 2A**). The shared pathways were mainly related to immune processes and the T1D positive control gene set containing top T1D hits from the GWAS catalog. Unique pathways for Cohort 1 included Influenza/HIV infection, nuclear envelope breakdown, transport of mature mRNA. Unique pathways for Cohort 2 included purine metabolism, inositol phosphate metabolism, and RNA degradation.

**Figure 2:**
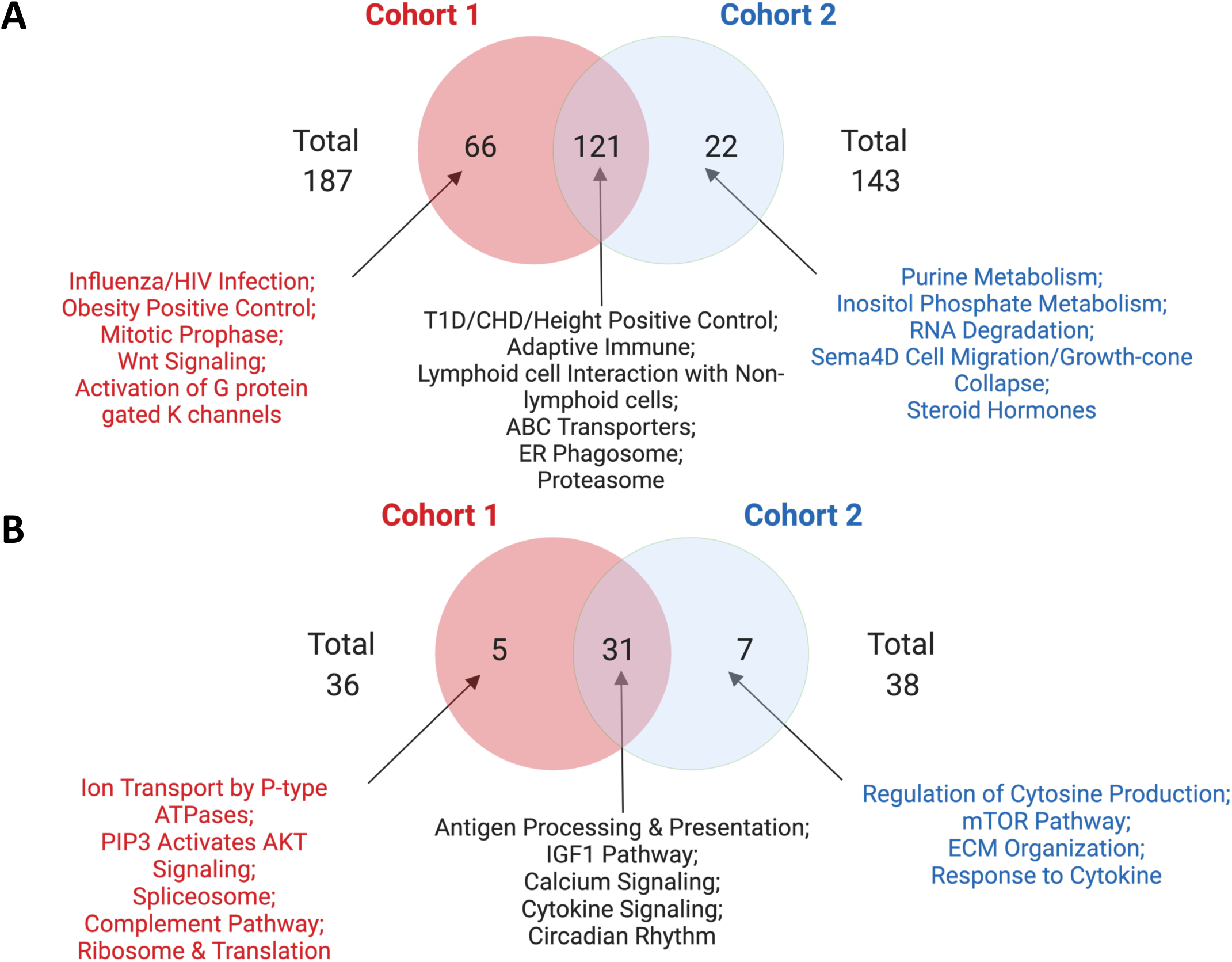
Venn Diagram of enriched canonical pathways and co-expression modules for both T1D GWAS cohorts. A) The independent and overlapping knowledge-driven biological pathways for both cohorts (FDR <5%). B) The independent and overlapping co-expression modules for both cohorts (FDR <5%).

Using WGCNA (49), we generated 272 data-driven gene coexpression modules from individual tissues from the GTEx transcriptomic database (44), with which we integrated our Cohort 1 and Cohort 2 T1D GWAS data sets separately. Cohort 1 showed enrichment in a total of 36 unique modules (FDR < 5%) in at least one tissue (**Supplement Table 3A**), while Cohort 2 showed significance in 38 modules (FDR < 5%) (**Supplement Table 3B**). Between the two cohorts, we found 31 overlapping modules associated with immune pathways, such as interferon signaling and cytokine signaling. We also identified 5 unique modules from Cohort 1 (e.g., AKT signaling, complement pathway, and ion transport) and 7 unique modules from Cohort 2 (e.g., cytosine production/response, mTOR pathways, extracellular matrix or ECM organization) (**Figure 2B**).

In addition to the main findings from the seven tissues, we found largely confirmatory results with the other tissue types tested across both cohorts, with adrenal gland, liver, and spleen revealing largely immune related signals and pituitary gland showing significance in *Wnt* signaling and insulin receptor signaling (**Supplement Table 4A** for Cohort 1, **Supplement Table 4B** for Cohort 2).

### Merging of pathways and coexpression modules into independent supersets

We focused on the shared 152 significant gene sets uncovered in our meta-analysis between Cohort 1 and Cohort 2 (31 coexpression modules and 121 canonical pathways) as they reflected reproducible signals for T1D genetic association (**Figure 2A**, **Figure 2B**). As the coexpression modules and pathways were obtained from various sources, the gene sets had the potential to share a high number of overlapping gene members. We found that 92 out of the 152 significant gene sets shared gene members with at least one other gene set. To reduce the redundancy, we merged the 92 overlapping gene sets into 13 independent supersets (**Table 1**). Interestingly, canonical pathways tended to merge with canonical pathways, and coexpression modules tended to merge with coexpression modules, suggesting the two types of gene sets exhibited different biological properties. The supersets represented diverse biological pathways, including adaptive immune system, complement cascade, cell cycle, viral infection, protein folding, RNA Polymerase I/III, signaling by GPCR, signaling by NOTCH, activation of GABA B receptor, B-cell receptor (BCR) signaling, and tRNA aminoacylation. The other 60 non-overlapping gene sets were kept intact, producing a total of 73 non-overlapping supersets. After using our merging algorithm of similar pathways, we ran a second round of MSEA to confirm whether our merged modules retained significance for T1D association. From the 73 non-overlapping supersets, we confirmed 44 supersets that were derived from canonical pathways (**Figure 3A; Supplement Table 5**) and 29 from coexpression modules (**Figure 3B**) to show statistical significance in our combined Cohort 1 and Cohort 2 datasets. Across the studied tissues, immune and cell cycle related processes were consistently enriched, particularly in monocytes, macrophages, lymphocytes, blood, adipose, and pancreas.

**Figure 3:**
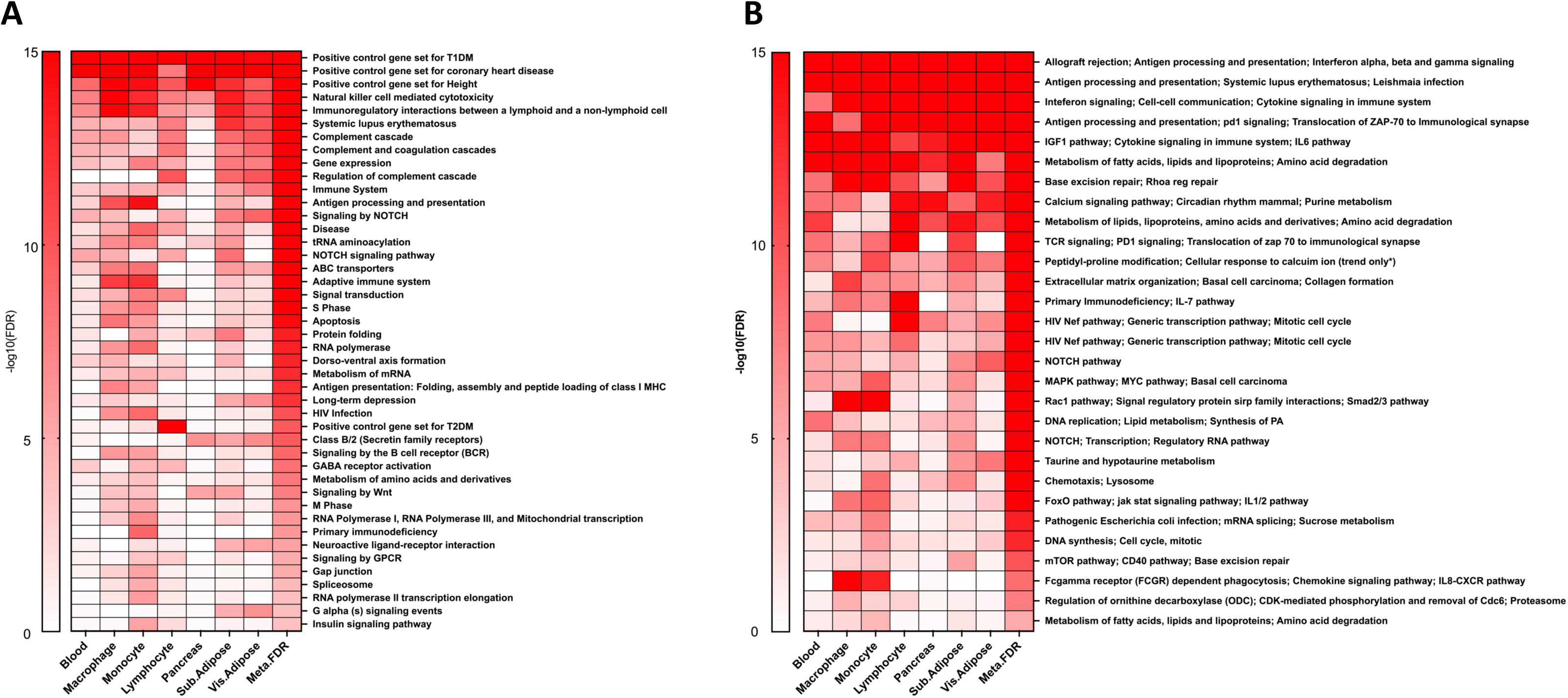
Heatmap of the tissue-specific meta-MSEA from the combined Cohort 1 and Cohort 2 datasets for the supersets derived from the canonical pathways and co-expression network modules. A) Heatmap for the statistical significance of T1D genetic association across the supersets derived from the Canonical pathways (FDR <5%) in the tissue-specific and cross-tissue analyses. B) Heatmap for the statistical significance of T1D genetic association across the supersets derived co-expression modules (FDR <5%) in the tissue-specific and cross-tissue analyses.

**Table 1.**
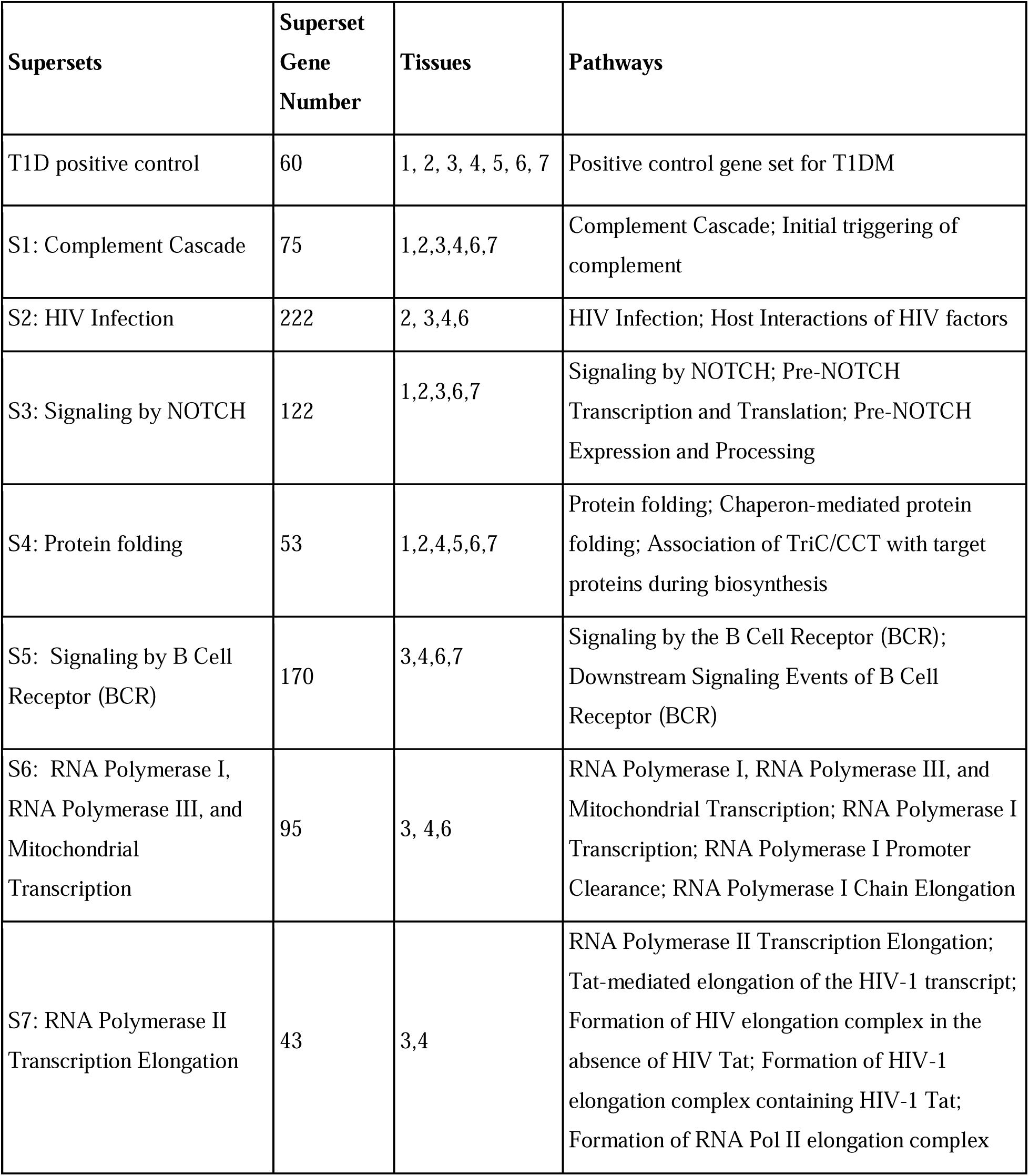
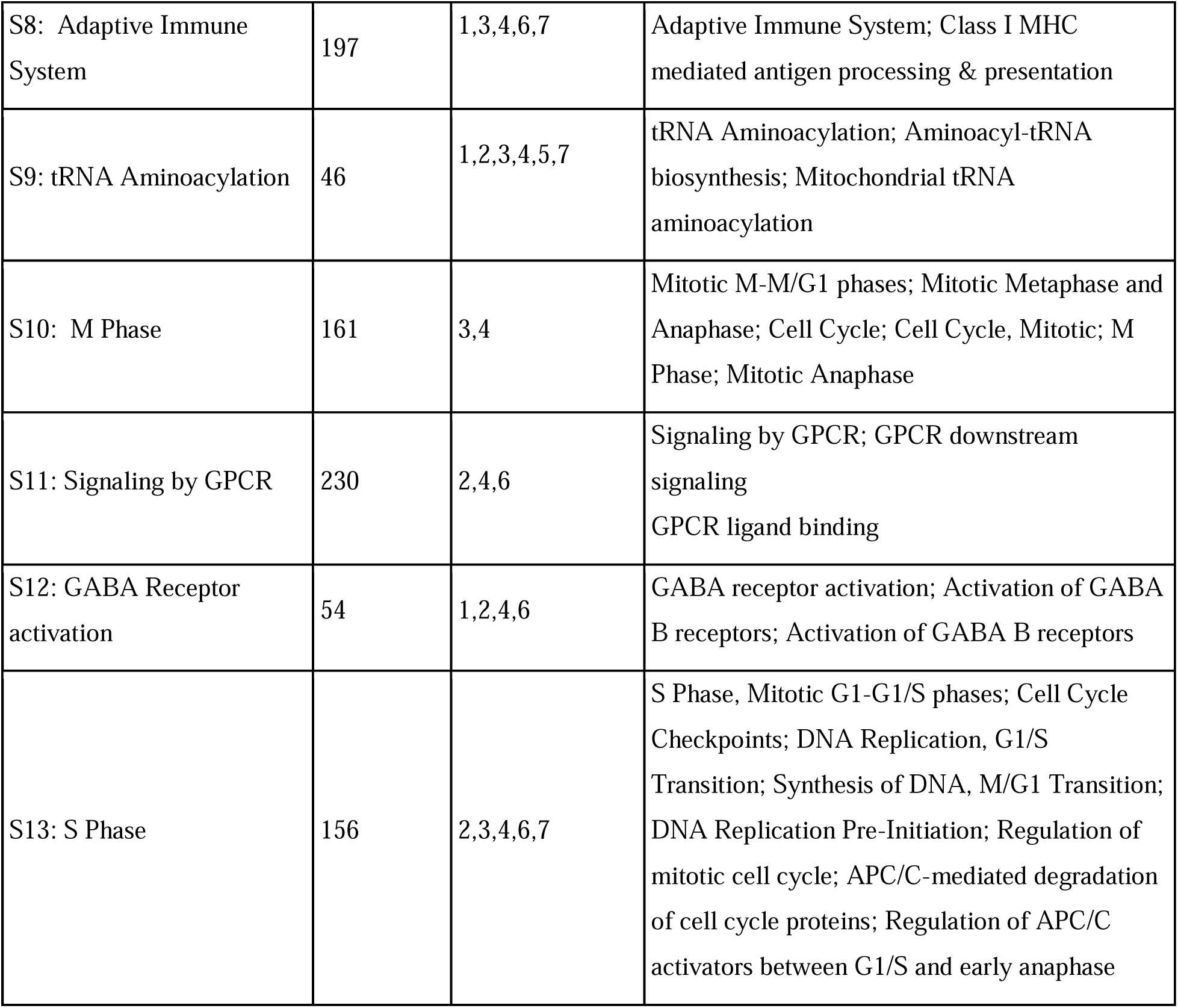
Top pathways associated with T1D identified across multiple tissues at an FDR <5%. 1: Blood, 2: Macrophage, 3: Monocyte, 4. Pancreas, 5: Subcutaneous Adipose, 6: Visceral Omentum Adipose, 7: Lymphocyte.

| **Supersets** | **Superset Gene Number** | **Tissues** | **Pathways** |
| --- | --- | --- | --- |
| T1D positive control | 60 | 1, 2, 3, 4, 5, 6, 7 | Positive control gene set for T1DM |
| S1: Complement Cascade | 75 | 1,2,3,4,6,7 | Complement Cascade; Initial triggering of complement |
| S2: HIV Infection | 222 | 2, 3,4,6 | HIV Infection; Host Interactions of HIV factors |
| S3: Signaling by NOTCH | 122 | 1,2,3,6,7 | Signaling by NOTCH; Pre-NOTCH Transcription and Translation; Pre-NOTCH Expression and Processing |
| S4: Protein folding | 53 | 1,2,4,5,6,7 | Protein folding; Chaperon-mediated protein folding; Association of TriC/CCT with target proteins during biosynthesis |
| S5: Signaling by B Cell Receptor (BCR) | 170 | 3,4,6,7 | Signaling by the B Cell Receptor (BCR); Downstream Signaling Events of B Cell Receptor (BCR) |
| S6: RNA Polymerase I, RNA Polymerase III, and Mitochondrial Transcription | 95 | 3, 4,6 | RNA Polymerase I, RNA Polymerase III, and Mitochondrial Transcription; RNA Polymerase I Transcription; RNA Polymerase I Promoter Clearance; RNA Polymerase I Chain Elongation |
| S7: RNA Polymerase II Transcription Elongation | 43 | 3,4 | RNA Polymerase II Transcription Elongation; Tat-mediated elongation of the HIV-1 transcript; Formation of HIV elongation complex in the absence of HIV Tat; Formation of HIV-1 elongation complex containing HIV-1 Tat; Formation of RNA Pol II elongation complex |
| S8: Adaptive Immune System | 197 | 1,3,4,6,7 | Adaptive Immune System; Class I MHC mediated antigen processing & presentation |
| S9: tRNA Aminoacylation | 46 | 1,2,3,4,5,7 | tRNA Aminoacylation; Aminoacyl-tRNA biosynthesis; Mitochondrial tRNA aminoacylation |
| S10: M Phase | 161 | 3,4 | Mitotic M-M/G1 phases; Mitotic Metaphase and Anaphase; Cell Cycle; Cell Cycle, Mitotic; M Phase; Mitotic Anaphase |
| S11: Signaling by GPCR | 230 | 2,4,6 | Signaling by GPCR; GPCR downstream signaling<br>GPCR ligand binding |
| S12: GABA Receptor activation | 54 | 1,2,4,6 | GABA receptor activation; Activation of GABA B receptors; Activation of GABA B receptors |
| S13: S Phase | 156 | 2,3,4,6,7 | S Phase, Mitotic G1-G1/S phases; Cell Cycle Checkpoints; DNA Replication, G1/S Transition; Synthesis of DNA, M/G1 Transition; DNA Replication Pre-Initiation; Regulation of mitotic cell cycle; APC/C-mediated degradation of cell cycle proteins; Regulation of APC/C activators between G1/S and early anaphase |

### Pathways and Coexpression Modules based on pancreatic islet eQTLs were highly consistent between the two cohorts

With the pancreatic islet being the primary site of destruction in T1D pathogenesis, we investigated this tissue region separately to capture the unique perturbed pathways. From the 1827 canonical pathways and the 272 coexpression modules we evaluated, we found a total of 8 pathways significant in Cohort 1 (**Supplement Table 6A**) and 16 pathways significant in Cohort 2 (**Supplement Table 6B**). The 8 pathways uncovered in Cohort 1 consisted of antigen presentation, calcium signaling, metabolism, transcriptional control and the T1D positive control gene set. All of these pathways were significant in Cohort 2 and displayed a stronger statistical significance. The 8 pathways unique to Cohort 2 include genes associated with coronary heart disease (CHD), immune pathways, height, and insulin receptor signaling. Given the smaller population size of Cohort 1 and the limited availability of islet eQTL expression data which reduce statistical power, we kept all 16 pathways uncovered in Cohort 2 (including the 8 pathways replicated in Cohort 1) in our downstream analysis (**Figure 4A**).

**Figure 4:**
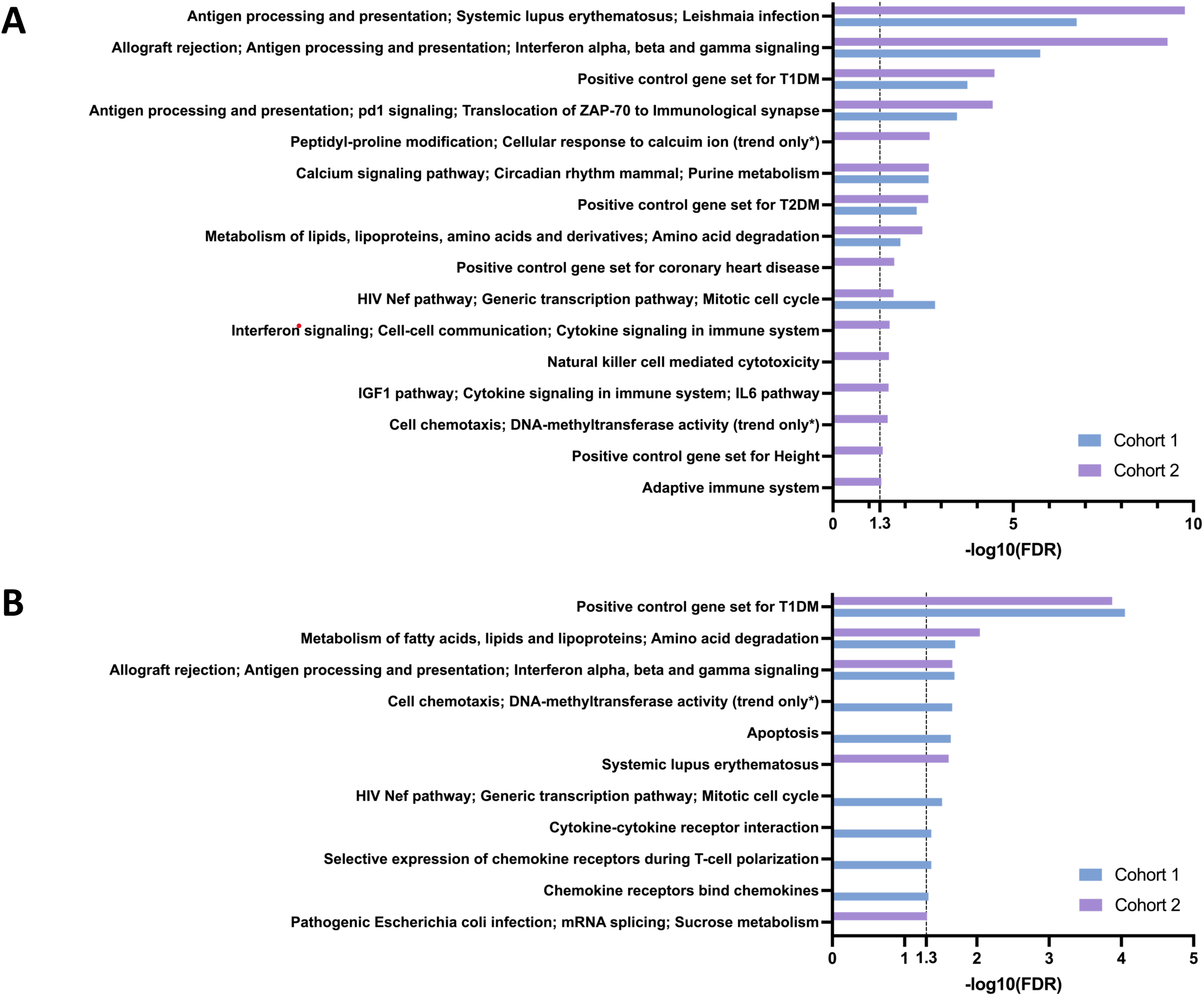
Bar graph for pancreatic islet MSEA results from the Cohort 1 and Cohort 2 datasets for the canonical pathways and co-expression network modules. Bar graph for statistical significance of the canonical pathways and co-expression modules (FDR < 0.05) in A) pancreatic islet tissue-specific analyses and B) pancreatic islet non-HLA tissue-specific analyses for each cohort independently.

### Removal of HLA genes from biological pathways and modules

As most of the known biology involved in T1D pathogenesis is concerned with the MHC region, specifically the class I and II (HLA) genes, we hypothesized that these genes may overshadow additional important processes involved in disease pathology and performed an additional MSEA analysis excluding the HLA genes from the 1872 canonical pathways and 272 coexpression modules.

We first examined the non-HLA MSEA results for pancreatic islets in Cohort 1 and Cohort 2 separately, identifying 9 significant pathways/modules in Cohort 1 (**Supplement Table 7A**) and 5 significant pathways/modules in Cohort 2 (**Supplement Table 7B**), with 3 overlapping ones between cohorts including the positive control gene set for T1D, metabolism of lipids, and antigen processing and presentation (**Figure 4B**). Other significant pathways in Cohort 1 included apoptosis, cell cycle, and immune related pathways (e.g., HIV pathway, cytokine receptor interaction, and chemokine receptor binding). The other two unique pathways in Cohort 2 are systematic lupus erythematosus and pathogenic *E. coli* infection.

We then focused our non-HLA MSEA analysis on the other seven informative tissues (blood, lymphocyte, macrophage, monocyte, pancreas, subcutaneous adipose, and visceral omentum adipose). We found a total of 166 gene sets to be significant (FDR<5%) in both Cohort 1 (**Supplement Table 7A**) and Cohort 2 (**Supplement Table 7B**), which were also significant in meta MSEA across both cohorts. After using our merging algorithm to reduce redundancy, we discovered a total of 64 non-overlapping supersets (**Supplement Table 8**). Comparing these supersets to the 73 non-overlapping supersets in our results from the HLA-inclusion analysis above, we noticed that 39 supersets were preserved when HLA genes were removed. These include immune pathways (e.g., adaptive immune system, cytokine signaling and signaling by B Cell receptors), insulin signaling, tRNA aminoacylation, apoptosis, viral infection, protein folding and complement cascade.

Importantly, removal of HLA genes uncovered 25 unique supersets absent in our HLA-inclusion results (**Supplement Table 8**). These supersets included immune related pathways such as antigen processing and RIG-I/MDA5 induction of IFN pathways, along with more diverse pathways including melanogenesis, steroid hormone biosynthesis, iron uptake and transport, and mitochondria in apoptotic signaling (**Figure 5**).

**Figure 5:**
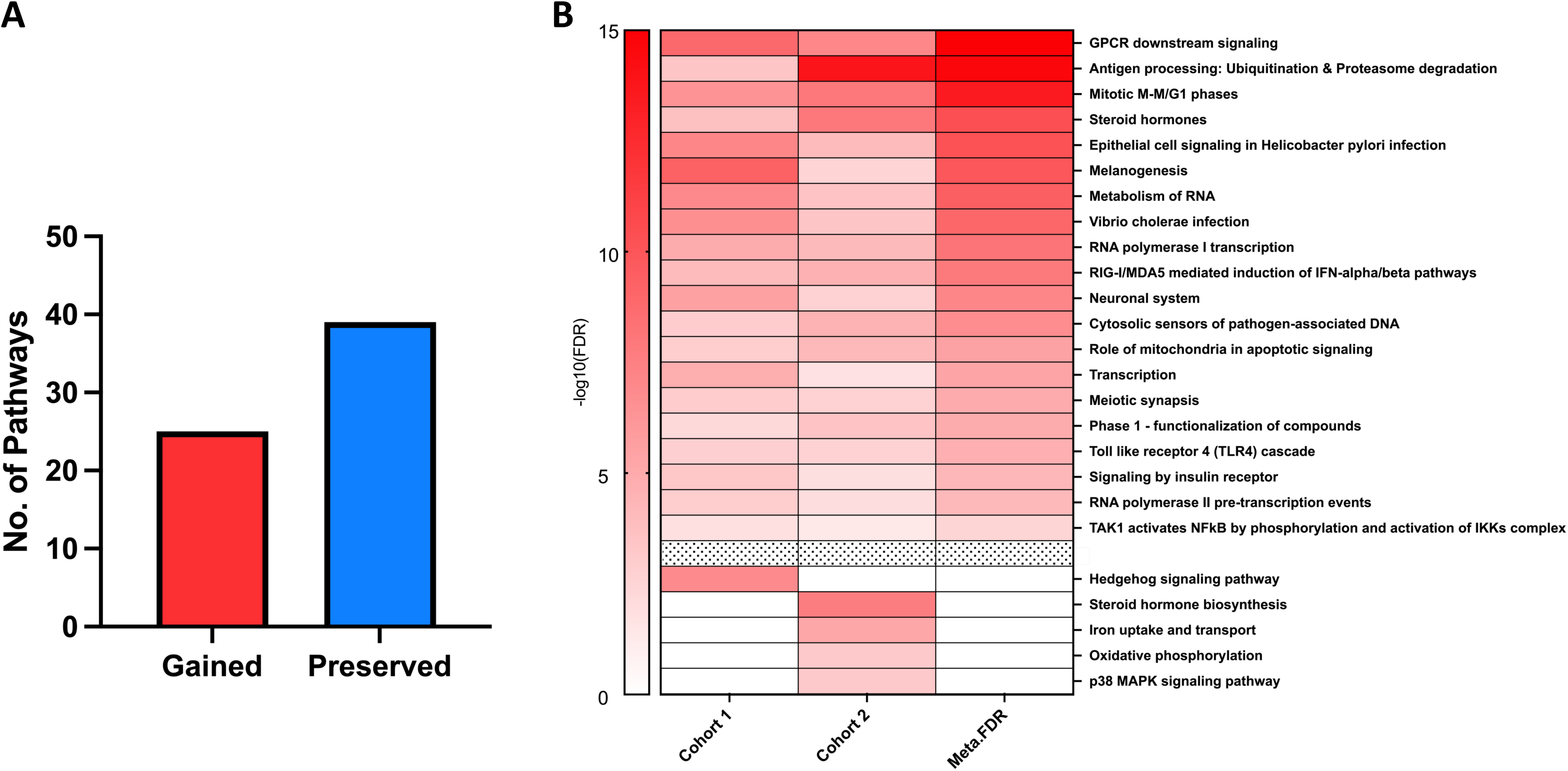
Graph and heatmap showcasing the number of preserved and gained pathways after removing the “HLA effect”. A) Bar graph denoting the number of gained versus preserved pathways (FDR<5%). B) Heatmap showing the gained pathways for both Cohort 1 and Cohort 2 above the black line (FDR<5%) and then the pathways that were significant (FDR<5%) in either Cohort 1 or Cohort 2 but not collectively.

### Identification of central regulators for T1D via a key driver analysis

To identify central regulatory genes, or key drivers (KDs) whose network neighbors are enriched for genes within the T1D associated supersets uncovered by MSEA, we performed a key driver analysis (KDA) on the T1D-associated gene sets uncovered above using protein-protein interaction (PPI) and tissue-specific Bayesian gene regulatory networks (GRNs) (53,54) (**Figure 6, Supplement Figure 1**).

**Figure 6:**
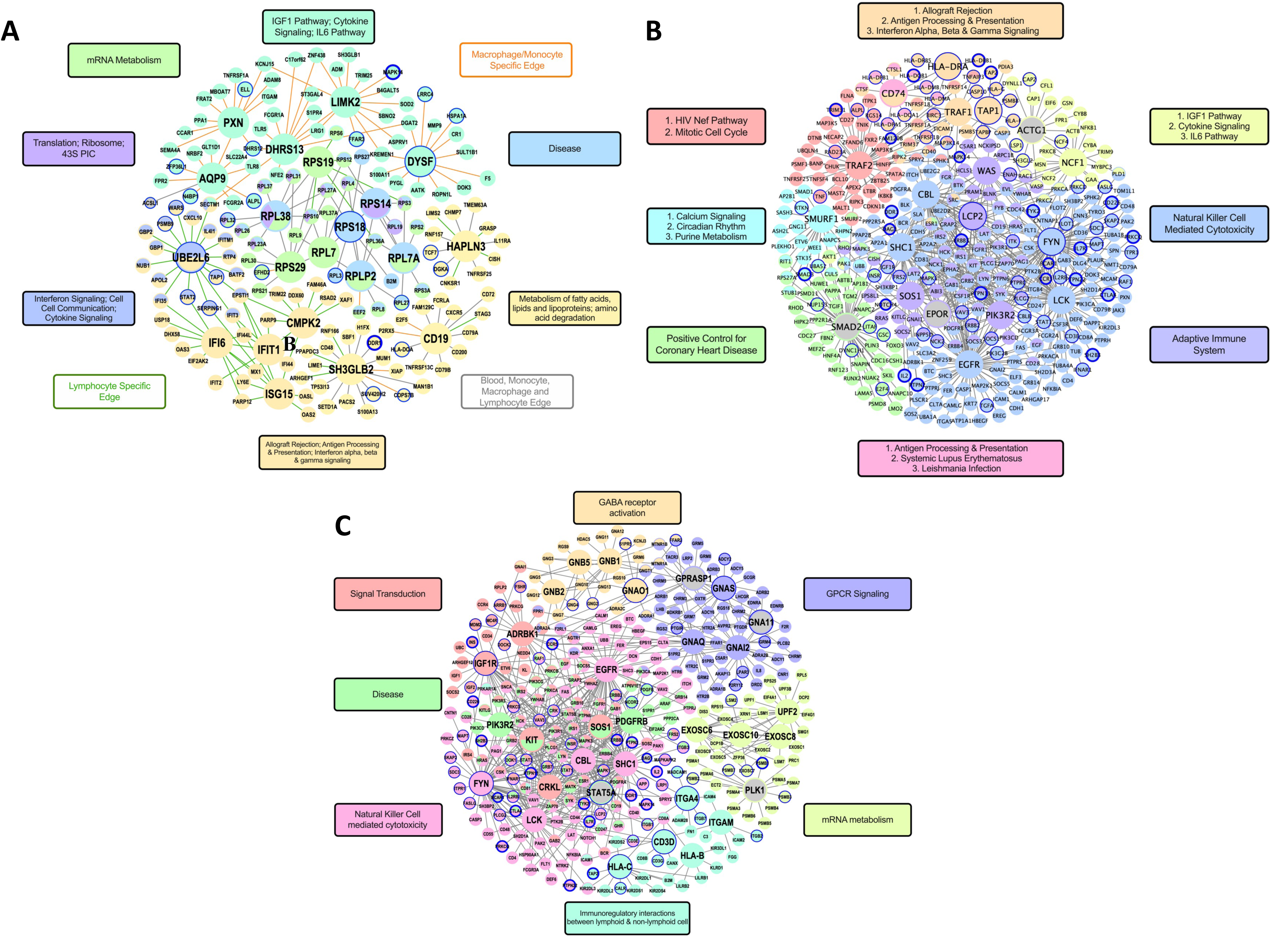
Bayesian Gene Regulatory Networks and PPI Networks. (A) Blood, macrophage, monocyte, and lymphocyte combined Bayesian network. (B) Islet PPI network. (C) Lymphocyte PPI Network.

#### Gene regulatory networks

In our islet Bayesian network, we utilized two approaches, one using islet eQTL mapped genes as input to our KDA and another using a combination of islet and hypothalamic eQTL (due to their similarity in expression patterns to the islet) mapped genes to increase power (55). We found that the islet specific network was sparser as expected but highlighted interesting KD candidates including *TREM2*, *LY86*, and *C1QC* under antigen processing and presentation and *PARP14* and *PSMB8*, which were related to interferon signaling. All of these KDs were also present in the combined hypothalamic/islet network but in addition we found KDs such as *CTSS* involved in inflammation and antigen presentation, *CFB* associated to the complement and coagulation cascade, and *RTP4* under antigen processing and presentation (**Supplement Figure 1A**).

Next, focusing on the most informative tissues outside of the islet, we found a total of 348 unique KDs from our Bayesian network across adipose, blood, macrophage, monocyte, lymphocyte, and pancreas networks at an FDR < 5% (**Supplement Table 9A**). To focus on the most central regulators, we chose the top five ranked KDs satisfying an FDR < 5% for each superset in each tissue-specific network. Among these, 17 were shared among at least 2 supersets and two KDs (*RPS29* and *RPS18)* were shared across the adipose, blood, lymphocyte, monocyte, and macrophage networks. Among the tissue networks, the adipose network revealed the largest number of 47 KDs (**Supplement Figure 1B**), of which 10 were shared with KDs uncovered in blood, lymphocyte, monocyte and macrophage networks (**Figure 6A**). The KDs uncovered are related to cell cycle, metabolism, and immune pathways. Notably, many of the KDs (e.g., *RPS29*, *RPLP2*, *CD19*, *IFIH1*, and *PTPN6*) have been found to be involved in viral infections, autoimmune and childhood onset disease, all of which are associated with T1D (56–58).

#### Protein-protein interaction (PPI) networks

To complement the results from using GRNs and to expand our search for KDs, we performed a KDA using the tissue-specific T1D pathways and PPI networks from the STRING database (59). Using the islet pathways, we identified top KDs such as *SMAD2*, *CD74*, *LCK*, *FYN*, *SHC1*, *EGFR*, *PIK3R2*, *TAP1*, and *HLA-DRA* (**Figure 6B, Supplement Table 9B**). For adipose, blood, lymphocyte, monocyte, macrophage and pancreatic tissues (**Supplement Table 9C**), we found a total of 134 KDs, with 7 specific to lymphocytes (*ITGAM*, *IGF1R*, *SOS1*, *CBL*, and *STAT5A*) (**Figure 6C**), 5 specific to adipose (*CASP9*, *FLNA*, *BRF1*, *CD74*, and *HLA-DRA*), 2 specific to blood (*PTPRC* and *MYC*), and 2 specific for macrophage (*POLR2G* and *PROS1*).

When comparing KDs from the PPI network with those from the Bayesian GRN networks, we find 26 overlapping KDs for tissue specific networks (24 for adipose, 2 consistent in blood and other tissues). The top 5 shared KDs (FDR<5%) from both types of networks are *LCK*, *VAV1*, and *PTPN6* for Natural Killer Cell Cytotoxicity as well as *F2* and *PLG* for Complement and Coagulation pathways. Other KDs include complement related genes such as *CD19* and *CD74*, DNA replication genes such as *MCM2* and *MCM6*, as well as those linked to autoimmunity such as *STAT1*, *SERPING1*, and *GZMB*.

#### Identification of Key Drivers of non-HLA specific supersets using PPI network

As the 20 additional pathways gained from the HLA-excluded analysis were found to be significant collectively across numerous tissues, we chose to perform a KDA using a PPI network rather than utilizing a tissue specific analysis through Bayesian networks (**Supplement Figure 1C**). We found a total of 33 unique KDs, some of which have previously been linked to T1D including *CASP3* and *CASP9* as KDs for the Role of Mitochondria in Apoptotic Signaling pathway, as well as *TBP* for the Transcription pathway. Several KD genes have also been studied in relation to type 2 diabetes and insulin resistance, including *AKT1* and *FGFR1* for the Signaling by Insulin Receptor pathway, *CTNNB1* for the Hedgehog signaling and Melanogenesis pathway, *IKBKB* for RIG-I/MDA5 induction of IFN pathways, and *CHUK* for the Cytosolic Sensors of Pathogen-associated DNA pathway. Additional KDs of interest include *RNF11* for Antigen Processing along with *RAF1* and *EGFR* for Melanogenesis.

### *In silico* validation of the Islet Key Drivers

To cross-validate the KDs for specific relevance to T1D, we examined the islet RNA sequencing and proteomics profiles from the Attie Lab Diabetes Database, where eight founder strains of mice, such as the B6 strain and the NOD mouse (a T1D model) were profiled (60). Among the islet PPI KDs, most were upregulated in NOD mice compared to the other seven non-diabetic mouse strains (**Figure 7A-F, Supplement Figure 2A-F**), except *SOS1* which showed downregulation (**Figure 7G**). Additionally, in female NOD mice, which are more susceptible to T1D, KD genes such as *WAS, TRAF1, LCP2, FYN, LCK, CD74*, and others showed higher gene expression in islet profiles. Furthermore, at the protein level KDs CD74 and TAP1 exhibited significantly higher protein expression in NOD mice compared to the other non-diabetic strains (**Figure 7H, I**). Within the islet Bayesian network, a majority of the 12 KDs, including *CTSS*, *MPEG1*, *PSMB8*, *C1QB*, *FSCN1*, and *GPB4*, were found to be significantly upregulated in NOD mice (**Supplement Figure 2G-L**). Therefore, the specificity of the network KDs to T1D was confirmed in independent mouse model data.

**Figure 7:**
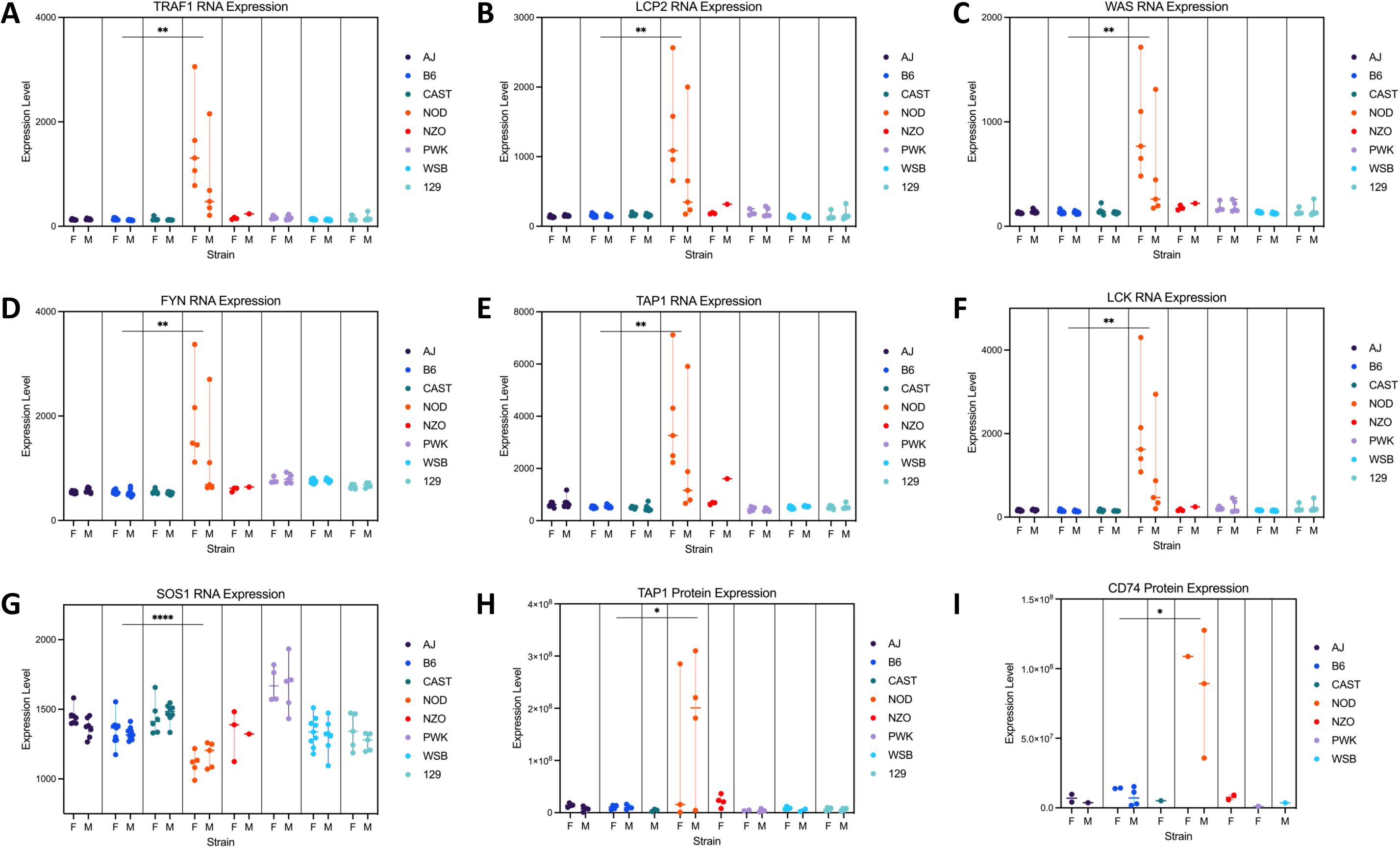
Example of in silico validation by screening for key driver gene RNA expression and proteomics patterns across seven non-type 1 diabetic mice with the type 1 diabetic NOD mouse. (A) RNA expression level of islet PPI KDs *TRAF1*, *LCP2*, *WAS*, *FYN*, *TAP1*, and *LCK* is significantly higher in NOD mouse. (B) RNA expression level of islet PPI KD *SOS1* is significantly lower in NOD mouse. (C) Protein expression level of islet PPI KD *CD74* and *TAP1* is significantly higher. (D) RNA expression level of islet Bayesian network KDs *PSMB8*, *MPEG1*, *GBP4*, *FSCN1*, *CTSS*, and *C1QB* is significantly higher in NOD mouse. p<0.05: *; p<0.01: **; p<0.001: ***; p<0.0001:****.

### Drug Repositioning

Utilizing our KD genes from individual tissues as an input into the drug repositioning tool LINCS 1000 as well as PharmOmics (61,62), we identified drugs that could be repositioned to target gene networks associated with T1D. For results from LINCS 1000, we filtered the drugs that pass a median Tau threshold of +/-90 as we cannot infer directionality from our input genes (**Figure 8A; Supplement Table 10**). From PharmOmics, we selected the significant drugs that ranked within the top 100 in each tissue and appeared at least five times in any of the screened tissues (**Figure 8B; Supplement Table 11**). We found of particular interest three main categories of drugs: Histone deacetylase (HDAC) inhibitors, Glycogen synthase kinase-3β (GSK-3β) inhibitors, and IκB Kinase (IKK) inhibitors.

**Figure 8:**
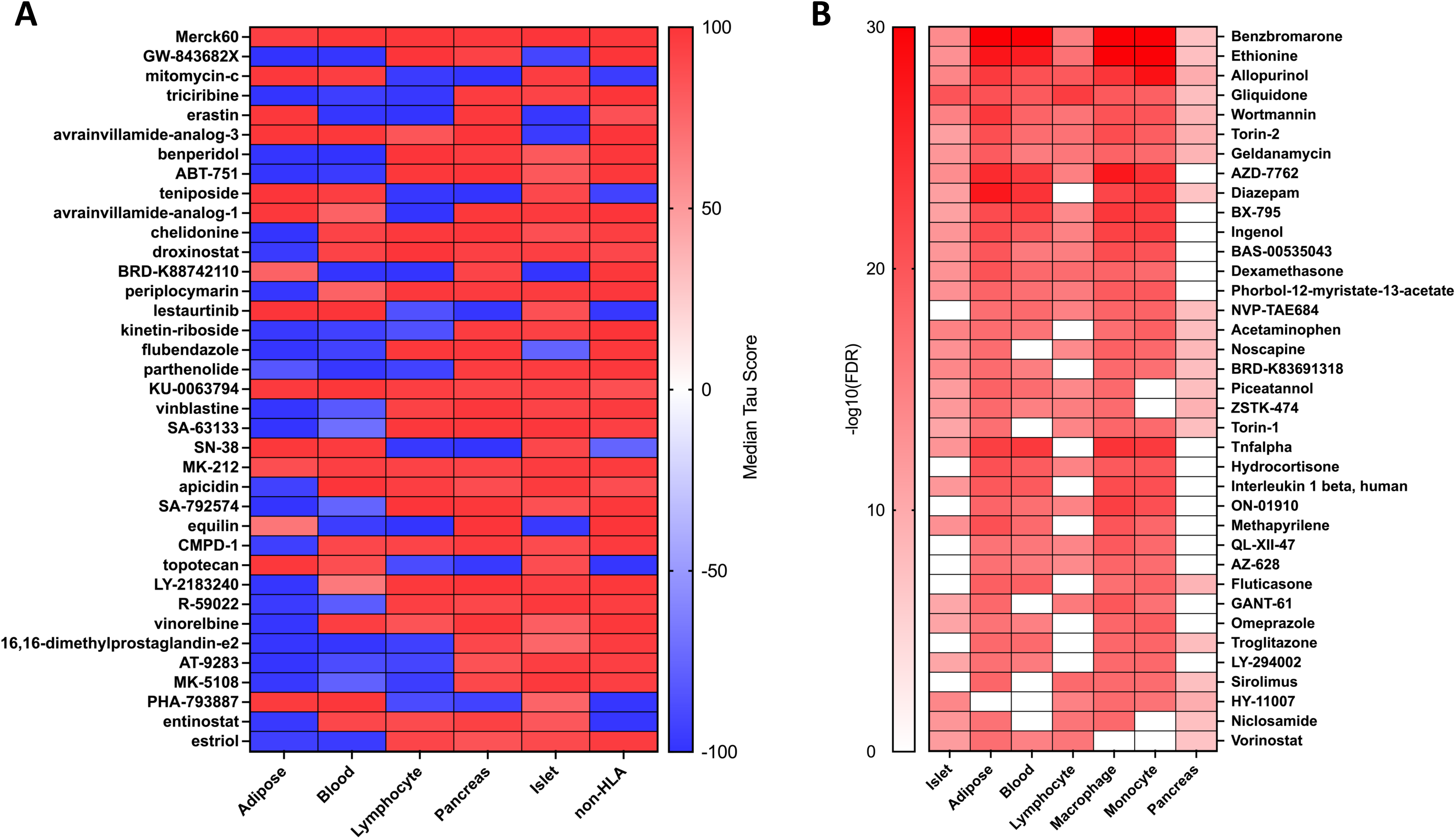
Drug repositioning results. A) Heatmap showcasing the top drugs repositioning using LINCS 1000 based on the key driver genes of each tissue/analysis. All Drugs collectively pass a Median Tau Threshold of 90 using the absolute mean across all groups. The highest median Tau score is presented in order from top to bottom. B) Heatmap of the top drug repositioning using Pharmomics based on key driver genes of each tissue. Only top significant 100 drugs were included for each tissue, and common drugs with more than 5 appearances across all tissues were demonstrated in heatmap.

First, for HDAC inhibitors, we found Apicidin, Etinostat, Merck60, Droxinostat, and Vorinostat, all selective inhibitors mainly targeting HDAC1, HDAC3, HDAC6, and HDAC9 to be top hits in our drug search across various tissues in both our PPI and Bayesian T1D networks.

Second, GSK-3β inhibitors, including kenpaullone and indirubin, were significant hits in our lymphocyte and pancreas networks, which agrees with previous literature that inhibition of GSK-3β showed protection against pancreatic beta-cell death (63). Kenpaullone is not only involved in GSK-3β inhibition, but also is a CDK inhibitor as well as a Src inhibitor, which may provide further protection to beta cells. Additionally, mTOR inhibitor Sirolimus and PI3K inhibitor LY-294002, which were significant drugs using adipose, macrophage, and monocyte network KDs, also indirectly regulate GSK-3β via the PI3K-AKT-mTOR signaling axis (64).

Finally, IKK inhibitors (TPCA-1, BMS-345541, BX-795, parthenolide, and IKK2-Inhibitor-V) were the third major drug class identified in our analysis of islet, lymphocyte, adipose, non-HLA and blood network KDs. IKK inhibitors have been found to be important targets in various autoimmune diseases such as rheumatoid arthritis, lupus erythematosus, multiple sclerosis, and irritable bowel disease, through their modulation of NFkB signaling, which is also known to be a key pathway in T1D (65).

Drugs of interest that only targeted the islet network KDs included tenofovir, PKCβ - inhibitors, and YM-976. All of these have been indicated for treatment of various immune and autoimmune diseases. Reverse transcriptase inhibitor tenofovir, administered as part of a Truvada tablet (HIV medication), was shown to improve hereditary autoimmune inflammation thought to be caused by retroelement cDNA buildup in mice (66).

Levonorgestrel, a progesterone receptor agonist and glucocorticoid receptor antagonist, was found to match with our lymphocyte key drivers. Increasing progesterone levels has been found to be protective against rheumatoid arthritis and multiple sclerosis; reducing stress hormones has been shown to reduce inflammation (67). Additionally, the top two hits in PharmOmics analysis, Benzbromarone and Allopurinol, play roles in reducing uric acid and lowering incidence of diabetes mellitus (68,69).

## DISCUSSION

Previous genetic studies have uncovered at least 60 loci linked with T1D development, yet our understanding of the intricate mechanisms underlying these associations and GWAS-based intervention approaches remain limited. To this end, we utilized a multi-omics integrative approach to advance our understanding of T1D etiology and prioritize potential therapeutic targets among the large number of disease-associated signals. The dysregulated pathways and key regulators of T1D etiology were elucidated by the combined analysis of the full summary statistics of T1D GWAS datasets (not just top genome-wide significant signals as typically studied), functional genomics data (tissue-specific eQTLs), knowledge-driven pathways, and data-driven gene coexpression and regulatory networks.

Our multiomics computational strategy confirmed known genes and biological pathways in T1D and revealed novel genes and their corresponding networks and biological processes. We identified a significant number of networks involved in immune and apoptosis related processes which have previously been implicated in T1D (70). We also found several pathways previously less associated with the genetic contribution of T1D such as viral infection, NOTCH signaling, amino acid degradation, and IGF-1 and insulin signaling pathways (71–73). Our tissue specific analyses revealed a consistent enrichment of antigen processing/presentation, natural killer cell cytotoxicity, and IFN-α/β/γ signaling across multiple tissues, supporting the immune origin of T1D highlighted in literature and emphasizing the presence of systemic dysregulation of immune function beyond insulitis (74).

As mentioned, beyond the classic immune components, we identified several pathways which imply a pathogenic change in the molecular machinery coordinating protein production and processing, including spliceosome in monocytes, mRNA metabolism in blood, macrophage, and monocytes, tRNA aminoacylation in macrophage, monocytes, and subcutaneous adipose, and proteasome in blood, macrophage, and monocytes. Kracht et al. reported the production of non-conventional products due to mRNA processing errors results in a polypeptide that is detected by T cells and generates an autoimmune response in T1D patients (75). It is also predicted that the formation of hybrid insulin peptides within beta cells activates CD4 T cells in NOD mice (76), thus further supporting the association between T1D and the failure of proteins to be processed correctly. It is plausible that variations within genes governing mRNA processing as well as protein formation and processing components induce antigenic protein products within the pancreatic tissue itself or within immune cells, which results in the activation of an autoimmune response, beta cell death, and development of T1D.

Importantly, within our tissue specific networks we were able to capture and highlight numerous previously implicated contributors to T1D, either as hub genes such as *IFIH1*, *HLA-C*, *SLC15A3*, *RAD51* (77) or peripheral genes *CTLA4*, *PTPN22*, *INS*, *INSR*, *HLA-DQA*. Of note, within the expected T1D immune pathways such as cytokine signaling, we uncover KD genes that are not well recognized for their specific role in T1D but consistent across tissues and are connected to well-known T1D GWAS hits. For example, *FYN,* a KD in both our lymphocyte (**Figure 6C**) and islet PPI networks (**Figure 6B**), was connected to important T1D associated GWAS loci (*CTLA4*, *INSR*, *CD226*, *PTPN11*). Furthermore, our *in-silico* validation showcased that *FYN* is upregulated uniquely in the NOD mouse model (**Figure 7**). This gene is essential in T cell signaling and interacts with *ZAP-70* (78) and *VAV1* (79,80), both of which are key components of T cell mediated immune response and are connected to *FYN* in our lymphocyte and islet PPI networks. In a similar fashion, we also highlight the KD *LCK* innetwork, which has also been previously shown to inhibit autoimmune responses through its interaction with *DUSP22* as a negative regulator of T-cell activation (81). *LCK* showed increased expression specifically in NOD mouse islets compared to non-T1D mouse models (**Figure 7**).

Interestingly, in the less well studied tissues for T1D such as adipose, we were able to capture previously documented T1D genes such as *SLC15A3* and *IFIH1. SLC15A3* is a T1D GWAS hit but limited knowledge exists for in its mechanistic connection to T1D pathogenesis. Our network analysis suggests a role of *SLC15A3* in signal transduction in adipose, including the regulation of inflammatory signals (82). Additionally, *IFIH1*, known to play a role in the innate immune response, is triggered by viral infections and has been shown to be a regulator of the diabetogenic T-cell response (83).

Viral infection (84) and its association with T1D have been suggested as the potential causal environmental trigger, particularly regarding antenatal maternal infection and subsequent incidence of T1D. In additional support of this, we found several pathways associated with HIV and influenza virus infection across multiple tissues tested. Given that these pathways are genetically perturbed as informed by our T1D GWAS datasets, our finding implies that genetic variants in genes involved in viral infection may confer vulnerability to infections and/or promote over-reactive viral response that induces autoimmunity, which may explain how viral infection trigger T1D pathogenesis. Moreover, we also find numerous KD genes known for their link with viruses but with limited evidence for direct genetic association with T1D such as *IFIT1*, which has been shown to be induced in NOD mice after rotavirus infection (85); *OAS2*, an innate immune activated antiviral enzyme (86); and *ISG15*, an antiviral effector linked with an anti-apoptotic effect in MIN6 cells (87). These genes were found to be within our blood and lymphocyte GRNs under the biological process interferon signaling and were key drivers in our T1D adipose tissue network. Additional KDs identified in our network analysis that are involved in susceptibility to viral perturbation include *CD3G* in adipose tissues (88), *PIK3R2* in lymphocytes and islets (89), and *IFI6* in lymphocytes.

To capture additional biology and KDs that may have been overshadowed by the “HLA effect”, we ran our pathway and network analysis after removing HLA-related genes. We recapitulated many key immune related processes showcasing the overarching and fundamental contribution of the immune system; these processes included antigen processing, NFkB and TLR4 cascades. More importantly, by removing HLA genes, we captured many pathways that may contribute to T1D through gene-environment interactions, such as viral infection pathways including RIG-I/MDA5 induction, *IFIH1-*related genes, and cytosolic sensors of pathogen-associated DNA; bacterial infection pathways such as epithelial cell signaling in *H. pylori* infection; melanogenesis, which may provide the link with a lack of Vitamin D and T1D development; and insulin receptor signaling which may interact with any or mixture of the above factors.

Our *in silico* validation of islet KDs highlights their significant role in the NOD mouse model of T1D. The upregulation of key drivers like *CTSS*, which degrades antigenic proteins (90), *CFB*, which contributes to complement activation (91), and *PSMB8*, induced by gamma interferon (92), underscores their potential involvement in T1D pathogenesis. The observed gender-specific expression, especially in female NOD mice, further supports the role of these KDs in the heightened susceptibility to T1D development in females in this model. The elevated protein expression of immune-related KD genes CD74 and TAP1 in NOD mice compared to other strains could indicate a more pronounced immune response, which aligns with the autoimmune nature of T1D. These findings contribute to a deeper understanding of the molecular underpinnings of T1D and may inform future therapeutic strategies.

Collectively taking into account all the T1D associated pathways and their corresponding networks, KDs, and tissues, we next aimed to find compounds that could be utilized to target these dysfunctional gene networks using the LINCS 1000 drug repositioning tool and PharmOmics gene-network-based drug repositioning tool (61,62). This exploratory analysis revealed numerous potential drug categories such as IKK inhibitors, GSK-3β inhibitors, and HDAC inhibitors. Additional interesting drugs included Benzbromarone, Allopurinol, and Omeprazole. Among our predicted drugs, HDAC inhibitors have been investigated previously with respect to T1D, particularly suberoylanilide hydroxamic acid (SAHA) and Trichostatin A (TSA), which were shown to prevent cytokine-induced beta cell toxicity in both primary rat islets and INS-1 cells but failed to produce normal insulin release upon glucose stimulation (93). ITF2357, another HDAC inhibitor, was shown to increase islet cell viability, reduce apoptotic cells and increase insulin secretion in STZ-induced T1D mice (94). A second class of drugs among our predictions is GSK-3β inhibitors such as alsterpaullone, which have been found to increase beta cell viability in cell culture in a dose-dependent manner by increasing ATP levels and reducing caspase 3 activity, which is important in apoptosis (95). In addition, it has been found that a selective GSK-3β inhibitor lithium chloride increased ATP levels but couldn’t reduce caspase 3 activity in the presence of cytokines. Our drug predictions also find kenpaullone, which acts as both a CDK inhibitor and a GSK-3β inhibitor and also targets additional genes within our networks, including *LCK* (96,97). Finally, we predicted IKK inhibitors like BMS-345541 as potential therapeutic options. IKK complex plays a key role in activating the NFkB pathway, and inhibition of this complex enhances beta-cell regeneration and could potentially slow down the rate of beta cell death (98). Therefore, plausible drugs were derived from our T1D GWAS-based pathway and network analysis.

Overall, our multitissue multiomics integrative analysis recapitulates previously known pathways, processes and genes associated with T1D pathogenesis, primarily components of the immune system, confirming the validity of the approach. We additionally uncover novel pathways and key genes involving multiple tissues that potentially contribute to T1D development through genetic perturbations, including those that can interact with environmental factors such as viral infections. These findings offer comprehensive understanding of T1D pathogenesis based on genetic evidence and through an omnigenic network lens. The drugs and KDs prioritized through our comprehensive integrative analyses may serve as putative T1D targets for therapies, which if experimentally validated in future studies may help reduce or prevent T1D progression.

## METHODS

### Overview of study design

We utilized an integrative genomics approach that leverages multiple large-scale human genetic and genomic datasets to elucidate the genetic networks and regulators of T1D pathogenesis (**Figure 1**). The datasets utilized included T1D GWAS from two independent cohorts, tissue specific eQTLs from diverse human tissues or cell types, various network models including gene coexpression networks, Bayesian gene regulatory networks and protein-protein interaction (PPI) networks, and biological pathway information (detailed in subsequent sections). To address reproducibility, we ran the integrative analysis on each GWAS study independently and then focused on the findings that were consistent between the two cohorts. For each GWAS, we mapped the single nucleotide polymorphisms (SNPs) to genes using tissue/cell-specific eQTL data with Marker Dependency Filtering (MDF) (25,30). The use of eQTLs helped inform on the most likely genes affected by GWAS SNPs based on functional evidence. Next, we grouped the genes based on whether they belonged to the same biological pathways or showed coexpression, which indicated functional relevance, in data-driven gene coexpression networks. We then assessed which pathways or gene coexpression modules (a module contains genes that show coexpression patterns) demonstrated stronger genetic associations with T1D compared to randomly generated gene sets using Marker Set Enrichment Analysis (MSEA) (25,30). After carrying out the MSEA process for each T1D GWAS dataset, we subsequently used a Meta-MSEA to meta-analyze the two independent GWAS data sets to look for shared pathways/modules that showed significant T1D associations, which we further simplified into independent “supersets” to reduce redundancy between pathways/modules. Integrating these T1D supersets with gene regulatory networks (Bayesian) and protein-protein interaction networks, we carried out the weighted key driver analysis (wKDA) to identify key drivers (KDs), which are central network genes whose network neighborhoods are highly enriched for genes in the T1D pathways and coexpression modules. These KDs were then visualized in tissue-specific networks. Furthermore, we carried out *in silico* validation to determine if these key drivers are linked with T1D. Lastly, drug repositioning were done to predict the potential drugs for treating T1D using LINCS 1000 and PharmOmics.

### T1D GWAS datasets

The summary statistics of GWAS for T1D was obtained from the JDRF/Wellcome Diabetes and Inflammation Laboratory, University of Oxford (43,99).

The study is comprised of 5913 T1D individuals of European descent (43,99), among which 3983 were genotyped using Illumina HumanHap550v3 (550k) Infinium Beadchip from the UK GRID and 1930 T1D individuals genotyped using Affymetrix 500K from the WTCCC. There were a total 8828 Controls, with 3999 genotyped using Illumina HumanHap550v3 (550k) Infinium Beadchip from the 1958 Birth Cohort (1958BC), 1490 genotyped using Affymetrix 500K (1958BC), 1455 genotyped using Affymetrix 500k from the UK Blood Services (UKBS), and 1884 genotyped using Affymetrix 500K from a cohort of bipolar disorders.

Inclusion criteria for the UK GRID are: T1D diagnosed between 6 months to 16 years of age, insulin dependent for greater than 6 months, a UK resident and self-identified as white European (average diagnosis age = 7.8 years of age, SD = 3.9, 47% female). Inclusion criteria for the WTCCC are: T1D diagnosed less than 17 years of age, insulin dependent for greater than 6 months and self-identified white European (average diagnosis age = 7.2 years of age, SD = 3.8, 49% female). Control inclusion criteria for UKBS and 1958BC included being residents in the UK and self-identified white Europeans. For the Bipolar cohort, control individuals greater than 16 years old and resident in the UK and of European descent were included.

The above T1D and control individuals were partitioned into two independent cohorts based on matching genotyping platforms (i.e., Illumina or Affymetrix) between cases and controls. Cohort 1 (Affymetrix) was comprised of 1930 T1D patients and 4830 Controls. Cohort 2 (Illumina) was comprised of 3983 T1D patients and 3999 Controls.

SNP genotypes were imputed to ∼10 million SNPs (1000 Genomes Phase III) using IMPUTE2, and routine quality controls were conducted as described in Cooper et al. (43). Statistical association between SNPs and T1D was carried out using a Bayesian analysis. All statistical association p-values for all imputed SNPs that passed quality control, regardless of significance level for T1D association, were used in our downstream analyses.

### Mapping SNPs to genes

To link GWAS SNPs to their potential target genes, tissue-specific eQTLs were used as they can provide functional insight for the role of SNPs in gene expression regulation within a given tissue. Thirteen eQTL sets were obtained from the GTEx database including subcutaneous adipose, visceral omentum adipose, blood, brain, colon, heart, liver, lymphocyte, muscle, pancreas, pituitary, spleen, and stomach (44,45). Additionally, we obtained macrophage and monocyte eQTLs from the Cardiogenics Consortium (29), pancreatic islet eQTLs from various sources (5), and immune cell eQTLs including lymphocytes from the DICE study (46). A broader spectrum of tissues was considered at this step to help objectively infer which tissues might be more informative for T1D association. GWAS was mapped to each tissue eQTL set separately to derive individual SNP-gene mapping sets reflecting tissue origins to allow assessment of tissue-specific signals.

A high degree of linkage disequilibrium (LD) was observed in the eQTL data, which may cause biases in the downstream analysis. For this reason, we removed redundant SNPs that had LD of r^2^ >0.7 with a chosen SNP. Briefly, a GWAS SNP was compared against other SNPs for LD and T1D association. If the SNP was in LD of r^2^ >0.7 with other SNPs, the one with the strongest T1D associations was chosen. This process was repeated until all remaining SNPs were not in LD based on the r^2^ >0.7 cut-off. These non-redundant SNPs were used for downstream analyses.

### Data-driven modules of co-expressed genes

In order to assess whether T1D GWAS signals are enriched in specific gene subnetworks, we derived gene coexpression networks using tissue-specific transcriptomic datasets from the GTEx portal, including subcutaneous adipose, visceral omentum adipose, blood, and pancreas (25,30,44). These tissues were chosen due to their relevance to T1D (33,34,36). The WGCNA (Weighted Gene Correlation Network Analysis) package was used to reconstruct coexpression networks based on gene expression profiles (49). Each tissue network contains multiple “modules”, and each module is comprised of tens to hundreds or thousands of genes that show coexpression. A total of 272 coexpression modules were curated.

### Knowledge-based biological pathways

We used a total of 1827 canonical pathways from Reactome (Version 45) (23), Biocarta (47), and the Kyoto Encyclopedia of Genes and Genomes (KEGG) databases (48). In addition to the knowledge-based pathways, we constructed a T1D positive control gene set based on candidate causal genes curated in GWAS catalog (p<5.0e-8) (100). Similar control gene sets for coronary heart disease (CHD), type 2 diabetes (T2D), and height were also constructed to compare with the T1D positive control set. Non-HLA specific pathways included all 1827 canonical pathways with HLA genes removed.

### Marker Set Enrichment Analysis (MSEA)

To identify coexpression modules and pathways that show evidence for genetic association with T1D, we applied MSEA from the Mergeomics package (25,30) on each of the GWAS cohorts separately in conjunction with the eQTL sources. MSEA employs a chi-square-like statistic with multiple quantile thresholds to assess whether a coexpression module or pathway shows enrichment of functional disease SNPs (i.e., those likely regulate gene expression as captured in eQTLs) compared to random chance. 10,000 permuted gene sets were generated for each coexpression module and pathway. As detailed in Shu et al., the enrichment statistics from the permutations were used to approximate a Gaussian distribution from which enrichment p-values were determined (30). Benjamini-Hochberg (BH) false discovery rate (FDR) was estimated across all coexpression modules and pathways tested for each GWAS. Gene sets were statistically significant if FDR < 5% in at least one SNP-gene mapping set. To evaluate gene sets across both GWAS studies, we followed up with a meta-analysis at the module/pathway level using the meta-MSEA function in Mergeomics, to retrieve robust gene sets across both cohorts. Stouffer’s Z score method was used to calculate meta p-values based on the p-values from the multiple MSEA runs. Meta-FDR was calculated using the Benjamini-Hochberg method, as described above.

### Merging overlapping pathways into supersets

The curated pathways and gene coexpression modules may carry redundant information. For example, a KEGG pathway “insulin signaling” can have largely overlapping genes with a Reactome pathway “insulin receptor signaling”. To reduce redundancy, we compared the significant modules and pathways associated with T1D at FDR <5% and merged the overlapping ones using a merging algorithm in Mergeomics to produce independent, non-overlapping “supersets” (25,30). The algorithm employs an overlap ratio *r* between two gene sets A and B as r = (r_AB_ x r_BA_)^0.5^, where r_AB_ is the proportion of genes in A that are also present in B and r_BA_ is the proportion of genes in B which are also in A. The overlap ratio cut-off was set to r >= 0.33 and Fisher’s exact test was used for assessing the statistical significance of gene overlap between modules/pathways. BH FDR < 5% was considered significant. Resultant supersets containing more than 500 genes were trimmed down to contain core genes shared among the overlapping gene sets.

### Tissue-specific gene regulatory networks and key driver analysis (KDA)

Tissue-specific Bayesian gene regulatory networks of adipose, blood, and pancreas tissue (25,30) as well as protein-protein interaction (PPI) networks were obtained from the Human Protein Reference Database (53). We chose to focus on these tissue networks due to our MSEA results showing the strongest statistical significance for these tissues. With these networks, we performed a key driver analysis using a KDA algorithm in Mergeomics to identify potential key drivers (KDs) whose network neighbors are enriched for genes within the T1D associated supersets uncovered by MSEA. The algorithm employed a chi-square like statistic similar to that described for MSEA, and FDR < 5% was used to focus on top robust KDs.

### *In silico* validation of KDs

To determine the relevance of the KDs to T1D, we examined the islet RNA sequencing and proteomics profiles from the Attie Lab Diabetes Database (http://diabetes.wisc.edu) (60). It is a searchable database with gene expression and proteomics profiles of different mouse strains, including NOD, a mouse model that spontaneously develops autoimmune diabetes. We then performed Student’s t-test on the RNA and protein expression levels for multiple genes between the NOD mice and B6 mice to look for significant differences.

### Drug Repositioning

We utilized the online web tool LINCS 1000 to match the best compounds and drugs based on the key driver genes of our given tissues and non-HLA related key drivers (62). We collected all compounds and drugs from each tissue and then calculated the absolute mean across blood (which includes macrophage and monocyte), lymphocyte, pancreas, pancreatic islet and non-HLA for the median tau score to list the drugs/compounds whose gene signatures best match each of our key driver networks.

We used PharmOmics web server for drug repurposing, focusing on the key drivers of the tissues, including islet, adipose, blood, lymphocyte, macrophage, monocyte, and pancreas (61). After obtaining the results, we filtered the drugs by significance (p-value < 0.05) and independently selected the top 100 drugs for each tissue. These selections were made according to how well the drug signatures matched the key drivers.

## Data Availability

All data produced in the present work are contained in the supplemental files

## ACKNOWLEDGEMENTS

We thank the Cardiogenics Consortium for providing eQTL resources, which were supported by the European Union FP6 program (LSHM-CT-2006-037593) and the Leducq CAD Genomics Transatlantic Network. We thank Dr. Jamie Inshaw and Dr. John Todd for providing the GWAS datasets.

## AUTHOR CONTRIBUTIONS

M.B., and X.Y., conceived and designed the research; M.B., Z.S., R.L., I.T., and J.W analyzed data. M.B., Z.S., and R.L., prepared figures; M.B., Z.S., R.L., J.W., and X.Y., drafted and edited manuscript; M.B., Z.S., R.L., I.H., J.W., and X.Y., approved final version of manuscript.

---

**Supplement Figure 1:**
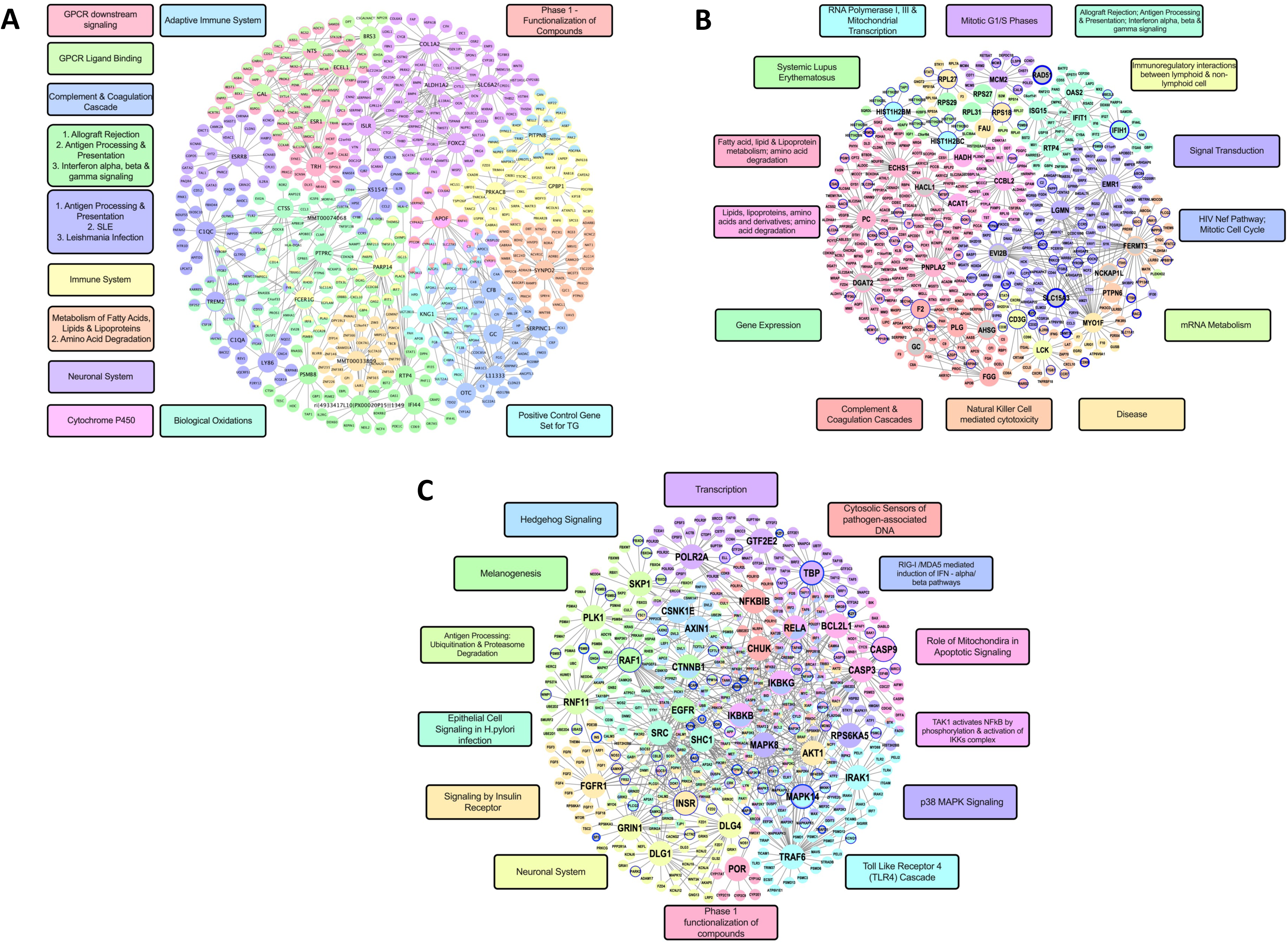
Bayesian Gene Regulatory Networks and PPI Networks. (A) Islet Bayesian network. (B) Adipose Bayesian network. (C) Non-HLA Network.

**Supplement Figure 2:**
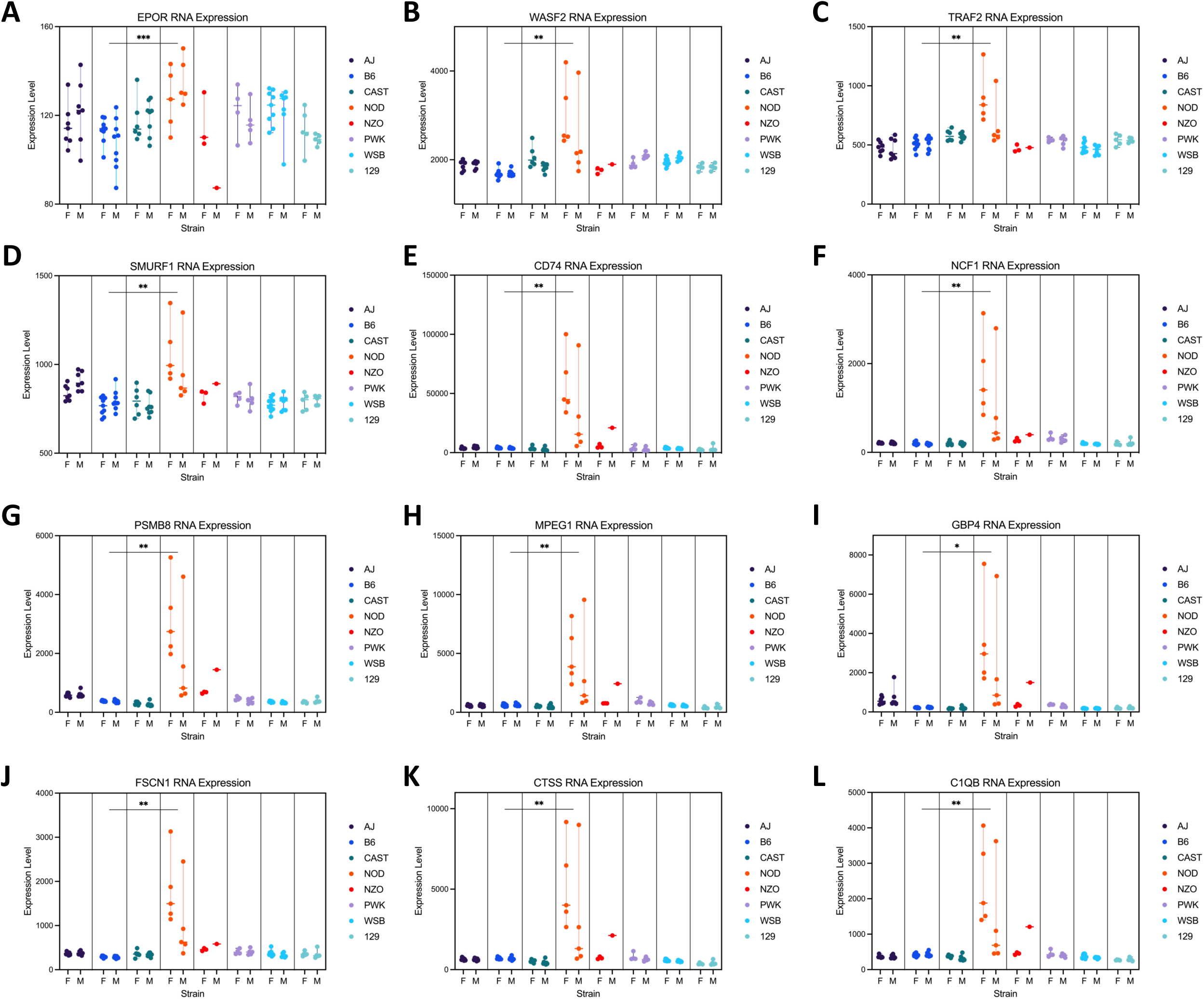
Example of in silico validation by screening for key driver gene RNA expression and proteomics patterns across seven nontype 1 diabetic mice with the type 1 diabetic NOD mouse. RNA expression levels of (A) *EPOR*, (B) *WASF2*, (C) *TRAF2*, (D) *SMURF1*, (E) *CD74*, (F) *NCF1*, (G) *PSMB8*, (H) *MPEG1*, (I) *GBP4*, (J) *FSCN1*, (K) *CTSS*, and (L) *C1QB* are significantly higher in NOD mouse. p<0.05: *; p<0.01: **; p<0.001: ***; p<0.0001:****

